# Plasma biomarkers, brain amyloid pathology, and cortical thickness in a diverse middle-aged community cohort: the HCP-CoBRA study

**DOI:** 10.1101/2025.07.10.25331312

**Authors:** Shayna T. Brodman, Nicholas Heaton, Gallen Triana-Baltzer, Xuemei Zeng, Alexandra Gogola, M. Ilyas Kamboh, Victor L. Villemagne, Oscar L. Lopez, Hartmuth Kolb, Rebecca A. Deek, Ann D. Cohen, Thomas K. Karikari

## Abstract

**INTRODUCTION:** We evaluated plasma biomarker association with, and classification accuracies for, Aβ-PET and cortical thickness in the biracial HCP-CoBRA cohort (53% B/AA and 47% NHW).

**METHODS:** In n=218 participants (age 62 [range: 57-71] years, 65% female and 15% Aβ PET-positive), plasma biomarkers (p-tau181, p-tau217, p-tau231, GFAP, NfL, Aβ42/Aβ40) were compared to Aβ-PET and MRI neuroimaging indicators.

**RESULTS:** P-tau217 (Janssen and ALZpath [AUCs=0.915-0.919]) had high sensitivity and specificity (>85%) for Aβ-PET status. All biomarkers except p-tau231 ruled out Aβ-pathology (NPV>95%) but only Janssen p-tau217+ was good for confirmation (PPV=0.909). Plasma biomarkers performed poorly for predicting cortical thickness but were elevated according to joint Aβ-PET-neurodegeneration profiles. Biomarker accuracies for Aβ-PET positivity were unaffected by self-identified race, except ALZpath p-tau217(p=0.024). However, correlations with Aβ-PET varied by self-identified race.

**DISCUSSION:** P-tau217 is a promising tool for Alzheimer’s disease-related Aβ pathology in older/middle-aged individuals. However, apparent race-related performances should be further studied.

## 1. BACKGROUND

Despite tremendous advancements in detecting and diagnosing Alzheimer’s disease (AD) according to cerebrospinal fluid (CSF) and neuroimaging techniques such as positron emission tomography (PET), these tests remain invasive, time-consuming, and expensive^1–5^. The development of plasma biomarkers for AD research has tremendous potential as screening and confirmatory diagnostic tools for use in intervention trials and clinical care programs^6–9^. The implications are vast including the potential for early detection with pre-clinical ramifications^7,10,11^. There is an abundance of literature regarding the accuracy and robustness of plasma biomarkers to detect neuropathological changes detected with brain imaging. However, studies are limited regarding community-based cohorts, specifically those comprised of diverse groups and backgrounds^11–14^. It is imperative to widen the scope of biomarker-focused investigations to communities and populations that are not routinely included in such studies, toward ensuring that clinically approved tests will factor in performance in a more representative group of participants.

The Amyloid-Tau-Neurodegeneration (AT(N)) framework proposed by the National Institute on Aging and the Alzheimer’s Association (NIA-AA) helps to standardize the diagnosis of AD based on the accumulation of amyloid-beta (Aβ; A), tau neurofibrillary tangles (T), and neuronal injury/degeneration (N) biomarkers^15,16^. Specifically, the biofluid-based biomarkers in the AT(N) scheme include CSF or plasma Aβ42/40 ratio, hyperphosphorylated tau (at various sites, including p-tau181, p-tau217 and p-tau231), and neurofilament (NfL) respectively^17^. Aβ42/40 ratio serves as a marker of amyloid plaque accumulation, typically altered early in preclinical stages^18^. Phosphorylated tau isoforms (e.g., p-tau181, p-tau217, p-tau231) are strongly associated with amyloid status and tau tangle formation, correlating with both disease progression and cognitive decline^19^. Neurofilament light chain (NfL) reflects neuronal injury and degeneration, though it is not specific to AD and may be elevated in other neurodegenerative disorders^20^. The recently updated AT(N) scheme recognizes the multifaceted pathophysiological processes in AD, expanding key processes of interest to include inflammation (I) and co-pathologies including vascular dysfunction (V) and concomitant neuropathologies including synucleinopathy (S)^21^. The inflammation marker glial fibrillary acidic protein (GFAP) is increasingly being recognized as a marker of astrocytic activation, which often co-occurs with amyloid pathology^22^. Together, these markers provide a multidimensional view of AD pathology and enable classification and staging. However, the impact of common social determinants of health, such as age, sex, *APOE* genotype, and, importantly, self-identified racial orientation, must be critically assessed for real-world application of these frameworks^23^. These factors might influence the measurements used in the diagnostic framework that use biomarkers to distinguish AT(N)^24^.

This study had a two-fold aim. Our first aim was to evaluate the clinical performance of plasma biomarkers in a racially diverse community cohort. We examined diagnostic accuracies of AT(N) plasma biomarkers (including p-tau181, p-tau217, p-tau231, Aβ40, Aβ42, GFAP, and NfL) to identify abnormal brain Aβ-PET scans as well as to identify abnormal cortical thickness. The second aim was to examine potential self-identified race-dependent association between these plasma biomarkers and Aβ PET in the same cohort.

## 2. METHODS

### 2.1 Participants

We included participants from the Human Connectome Project – Connectomics in Brain Aging (HCP-CoBRA), which recruited both cognitively normal and impaired participants, from Pittsburgh, PA, USA, aged 50-89 years^25^. Different sources of recruitment were employed including via the University of Pittsburgh Alzheimer’s Disease Research Center (Pitt-ADRC), the Pitt + Me web portal, which was primarily used for the recruitment of non-college educated Black/African American (B/AA) and non-Hispanic White (NHW) individuals, or through active links and word of mouth. The participants underwent both clinical and cognitive assessments over three days, beginning with an informed consent visit. Participants completed functional magnetic resonance imaging (fMRI) tasks both with fasting and then without fasting, as well as underwent magnetoencephalography (MEG) and ^11^C Pittsburgh Compound B (PiB) PET imaging for brain Aβ plaques a week after the last MRI scanning. Each participant also underwent a brief neuropsychological assessment based on the Pittsburgh ADRC protocol, including the Montreal cognitive assessment (MoCA), verbal fluency test, a 30-item visual naming test, Trailmaking, verbal free recall, and the Rey-Osterreith Complex Figure^25,26^. Demographic information, including self-identified racial identity, sex, and education, were also collected. HCP inclusion criteria and study design have been described in previous publications^25^.

All participants completed the informed consent process and intake forms. The HCP study and its protocols were reviewed and approved by the University of Pittsburgh Institutional Review Board.

### 2.2 Neuroimaging procedures

Imaging was conducted and processed as previously described^25^. Briefly, A status was based on a global [^11^C] PiB standardized uptake value ratio (SUVR) calculated by volume-weighted averaging of nine composite regional outcomes (anterior cingulate, posterior cingulate, insula, superior frontal cortex, orbitofrontal cortex, lateral temporal cortex, parietal, precuneus, and ventral striatum)^27^. Participants were classified as A+ or A-based on a pre-defined cutoff, with >1.346 as A+^28,29^. We classified N status based on MRI scans for cortical thickness (CT)^30–32^. N status was determined by an AD-signature composite CT index obtained from a surface-area weighted average of the mean CT of four FreeSurfer regions of interest (ROIs) – entorhinal cortex, inferior temporal and middle temporal gyri, and fusiform – all of which are most predictive of AD-specific pathology and diagnosis, with < 2.7 as N+^28,33^.

### 2.4 Plasma collection procedure

Blood collection and processing followed established protocols^34^. Blood samples were collected in 10 mL K2-EDTA tubes (BD Biosciences Cat# 366643) and centrifuged within 2 hours at 2000x*g* for 10 min at 4 °C to obtain the plasma portion of the whole blood. The plasma samples were then aliquoted and stored at −L80L°C until use. Buffy coats were also collected to determine the *APOE* genotype according to the published procedure^35^.

### 2.5 Procedures for Single molecule array (Simoa) assays

Simoa assays for the analytes p-tau181, p-tau217, p-tau231, Aβ40, Aβ42, GFAP, and NfL were performed on an HD-X (Quanterix, Billerica, MA, USA) as described by Zeng and colleagues^30^. Briefly, plasma samples were thawed at room temperature and centrifuged at 4000x*g* for 10 min to remove particulates. Plasma Aβ40, Aβ42, GFAP, and NfL were measured using the Neurology 4-Plex E (#103670) kit. P-tau217 was measured with both the ALZpath Simoa® p-Tau 217 V2 Assay (#104371) and the Janssen p-tau217+ assay, an in-house assay developed at Janssen Research & Development using Janssen’s proprietary antibodies^36^. P-tau181 was measured using the Karikari et al., method^37^, using p-tau181 antibody AT270 for capture and anti-Tau6-18 antibody (Tau12; BioLegend, # 806502) for detection. P-tau231 was measured in the same fashion but with p-tau231 antibody AT180 for capture and Tau12 for detection^37^. Quality control (QC) samples were analyzed at the start and the end of each run in 2-3 different concentrations per assay to assess the reproducibility of each assay. For p-tau181 and ALZpath p-tau217, the within- and between-plate coefficients of variation (CVs) were <5% and <6%, respectively. For the Neurology 4-plex assays, the within- and between-plate CVs were mostly <10% and <16%, respectively. The Janssen p-tau217+ assay had within- and between-plate CVs of 4.0 and 7.3%, respectively.

### 2.6 Statistical analysis

Continuous variables were reported as median (IQR) and categorical variables as counts (%). Statistical testing procedures to compare demographic characteristics and blood-based biomarkers according to A or N status were done via the Wilcoxon rank sum test for continuous variables and the Pearson’s Chi-squared test for categorical variables. Spearman’s correlations between biomarkers and continuous A or N status were also reported. Logistic regression models were fit to predict either A or N status. Univariable models for each biomarker were fit, as well as multivariable models with demographic and genetic characteristics, sex, age (at enrollment), and *APOE* ε4 carrier status. This allowed for the assessment of each biomarker’s predictive capabilities before and after adjustment for demographic variables. Any participants from the HCP cohort that had missing demographic characteristics were excluded from this study. A complete case analysis of all p-tau biomarkers was conducted first. Then, a reduced dataset was utilized for complete case analysis of NfL, GFAP, and Aβ42/40 ratio measured in a subset of samples with available volume for the Neurology 4-plex assay.

We trained and tested the models using a 50-50 training-testing data split, allowing the logistic regression models to be trained with 50% of the data and validated on the remaining 50%. This procedure was done for the prediction of both A and N status models. The area under the receiver operating characteristic curve (AUC) was calculated from the results of the testing dataset and reported with corresponding 95% confidence intervals^38^. We compared the biomarker-only models with the models that incorporated demographic variables via DeLong’s test for differences in AUC^38^.

For analysis where participants were grouped according to both A and N statuses, to create four possible groups, A-N-, A+N-, A-N+, and A+N+, all continuous variables were first compared using a Kruskal-Wallis rank sum test. For biomarkers with a significant global test, pairwise comparisons were made using Wilcoxon rank sum tests, with a Bonferroni correction for multiple comparisons adjustment. Demographic categorical variables were compared via either a Pearson’s Chi-squared or Fisher’s exact test.

Finally, participants were grouped by self-identified race, Black/African American (B/AA) or Non-Hispanic White (NHW), and by Aβ-PET status (A status). Biomarker assays were first compared between self-identified racial groups via Wilcoxon rank sum test. Statistical testing was also performed within each self-identified racial group, with continuous variables compared via Wilcoxon rank sum test and categorical variables with either Fisher’s exact test or Pearson’s Chi-squared test. Logistic regression models were fit using binary Aβ-PET status as the outcome variable. These models included one of the plasma biomarkers, self-identified race, and a term for their interaction to assess if the relationship between each biomarker and Aβ-PET status is altered by self-identified racial group. Similarly, linear regression models were fit to predict global PiB SUVR using the same predictors to assess if self-identified racial group alters the relationship between biomarker and global PiB SUVR. The plasma biomarker assays were standardized with the Z-transformation to make the interaction coefficients more comparable across the linear regression models.

## 3. RESULTS

### 3.1 Participant characteristics

We included 218 participants who had Aβ-PET (A status) data, including 185 (85%) and 33 (15%) A- and A+ individuals, respectively (see Table 1 for participant characteristics according to Aβ-PET abnormality). Among them, 116 (53%) were self-identified as B/AA, while 102 (47%) as NHW. The A+ participants had a median (IQR) age of 72 (68, 78) years, 14 (42%) female, 13 (39%) *APOE* ε4 carriers, and 8 (24%) self-identified as B/AA. The A-participants had a median (IQR) age of 60 (56, 67) years, 127 (69%) were female, 54 (29%) were *APOE* ε4 carriers, and 108 (58%) self-identified as B/AA (Table 1). Compared to the A- group, the A+ participants were older (p<0.001), had fewer females (p=0.004), and fewer participants self-identifying as B/AA (p<0.001), but no significant difference in the frequency of *APOE* ε4 carriers (p=0.242).

**Table 1.** Participant characteristics of the HCP cohort according to Aβ PET abnormality and plasma biomarker information.

| | Overall, n=218<sup>1</sup> | Aβ PET negative, n=185<sup>1</sup> | Aβ PET positive, n=33<sup>1</sup> | p-value<sup>2</sup> |
| --- | --- | --- | --- | --- |
| **Sex** |  |  |  | 0.004 |
| Female | 141 (65%) | 127 (69%) | 14 (42%) |  |
| Male | 77 (35%) | 58 (31%) | 19 (58%) |  |
| **Age at enrollment (years)** | 62 (57, 70) | 60 (56, 67) | 72 (68, 78) | <0.001 |
| **APOE4** |  |  |  | 0.242 |
| Non-carrier | 151 (69%) | 131 (71%) | 20 (61%) |  |
| Carrier | 67 (31%) | 54 (29%) | 13 (39%) |  |
| **Race** |  |  |  | <0.001 |
| Black/African American | 116 (53%) | 108 (58%) | 8 (24%) |  |
| Non-Hispanic White | 102 (47%) | 77 (42%) | 25 (76%) |  |
| **ALZpath p-tau217 (pg/mL)** | 0.25 (0.18, 0.36) | 0.24 (0.17, 0.31) | 0.51 (0.40, 0.80) | <0.001 |
| (Missing) | 4 | 3 | 1 |  |
| **Janssen p-tau217+ (pg/mL)** | 0.035 (0.026, 0.050) | 0.033 (0.025, 0.044) | 0.070 (0.049, 0.105) | <0.001 |
| (Missing) | 22 | 19 | 3 |  |
| **p-tau181 (pg/mL)** | 13 (9, 18) | 12 (9, 18) | 16 (12, 23) | 0.014 |
| (Missing) | 10 | 7 | 3 |  |
| **p-tau231 (pg/mL)** | 9 (6, 14) | 9 (6, 13) | 10 (6, 17) | 0.472 |
| (Missing) | 10 | 7 | 3 |  |
| **Aβ40 (pg/mL)** | 68 (53, 79) | 69 (53, 79) | 64 (54, 78) | 0.862 |
| (Missing) | 63 | 58 | 5 |  |
| **Aβ42 (pg/mL)** | 4.82 (3.71, 5.61) | 4.89 (3.90, 5.67) | 3.74 (3.30, 5.14) | 0.013 |
| (Missing) | 63 | 58 | 5 |  |
| **Aβ42/40 ratio** | 0.072 (0.061, 0.080) | 0.074 (0.065, 0.083) | 0.060 (0.056, 0.069) | <0.001 |
| (Missing) | 63 | 58 | 5 |  |
| **GFAP (pg/mL)** | 70 (49, 102) | 63 (46, 95) | 92 (79, 155) | <0.001 |
| (Missing) | 63 | 58 | 5 |  |
| **NfL (pg/mL)** | 13 (9, 18) | 12 (8, 17) | 18 (11, 22) | 0.003 |
| (Missing) | 63 | 58 | 5 |  |
| **Global PiB SUVR** | 1.11 (1.06, 1.19) | 1.09 (1.05, 1.14) | 1.66 (1.52, 1.91) | <0.001 |
| <sup>1</sup> n (%): Median (IQR) |  |  |  |  |
| <sup>2</sup> Pearson's Chi-squared test for categorical and Wilcoxon rank sum test for numerical variables |  |  |  |  |

The neurodegeneration status (N status) analysis included 220 participants with complete CT-based MRI data, with 142 (65%) and 78 (35%) being N- and N+, respectively (see Table 2 for demographic characteristics according to neurodegeneration status as assessed by cortical thickness). There was no significant difference between the N+ and N- groups in terms of age (p=0.814), sex (p=0.434), *APOE* ε4 carrier status (p=0.056), or self-identified race (0.119). The N+ participants had a median (IQR) age of 62 (56, 71) years old, 53 (68%) were female, 30 (38%) were *APOE* ε4 carriers, and 47 (60%) self-identified as B/AA. The N- participants had a median (IQR) age of 63 (57, 70) years, 89 (63%) were female, 37 (26%) were *APOE* ε4 carriers, and 70 (49%) self-identified as B/AA.

**Table 2.** Participant characteristics of the HCP cohort according to neurodegeneration status as assessed by cortical thickness.

|  | Overall, n=220<sup>1</sup> | Neurodegeneration-negative, n=142<sup>1</sup> | Neurodegeneration-positive, n=78<sup>1</sup> | p-value<sup>2</sup> |
| --- | --- | --- | --- | --- |
| **Sex** |  |  |  | 0.434 |
| Female | 142 (65%) | 89 (63%) | 53 (68%) |  |
| Male | 78 (35%) | 53 (37%) | 25 (32%) |  |
| **Age at Enrollment (years)** | 62 (57, 71) | 63 (57, 70) | 62 (56, 71) | 0.814 |
| **APOE4** |  |  |  | 0.056 |
| Not Carrier | 153 (70%) | 105 (74%) | 48 (62%) |  |
| Carrier | 67 (30%) | 37 (26%) | 30 (38%) |  |
| **Race** |  |  |  | 0.119 |
| Black/African American | 117 (53%) | 70 (49%) | 47 (60%) |  |
| Non-Hispanic White | 103 (47%) | 72 (51%) | 31 (40%) |  |
| **ALZpath p-tau217 (pg/mL)** | 0.25 (0.18, 0.37) | 0.24 (0.18, 0.34) | 0.27 (0.19, 0.43) | 0.109 |
| (Missing) | 4 | 2 | 2 |  |
| **Janssen p-tau217+ (pg/mL)** | 0.035 (0.026, 0.050) | 0.034 (0.026, 0.046) | 0.038 (0.025, 0.065) | 0.161 |
| (Missing) | 22 | 16 | 6 |  |
| **p-tau181 (pg/mL)** | 13 (9, 18) | 12 (8, 18) | 14 (11, 19) | 0.011 |
| (Missing) | 10 | 6 | 4 |  |
| **p-tau231 (pg/mL)** | 9 (6, 14) | 8 (6, 13) | 10 (7, 15) | 0.029 |
| (Missing) | 10 | 7 | 3 |  |
| **Aβ40 (pg/mL)** | 68 (53, 79) | 68 (53, 82) | 68 (53, 78) | >0.900 |
| (Missing) | 63 | 39 | 24 |  |
| **Aβ42 (pg/mL)** | 4.82 (3.71, 5.61) | 4.82 (3.73, 5.71) | 4.79 (3.60, 5.43) | 0.630 |
| (Missing) | 63 | 39 | 24 |  |
| **Aβ42/40 ratio** | 0.072 (0.061, 0.080) | 0.072 (0.062, 0.083) | 0.072 (0.061, 0.079) | 0.591 |
| (Missing) | 63 | 39 | 24 |  |
| **GFAP (pg/mL)** | 70 (49, 102) | 68 (49, 99) | 76 (55, 107) | 0.517 |
| (Missing) | 63 | 39 | 24 |  |
| **NfL (pg/mL)** | 13 (9, 18) | 13 (9, 18) | 15 (10, 21) | 0.281 |
| (Missing) | 63 | 39 | 24 |  |
| **CT Composite Region** | 2.74 (2.66, 2.79) | 2.78 (2.74, 2.82) | 2.63 (2.55, 2.67) | <0.001 |
| <sup>1</sup> n (%): Median (IQR) |  |  |  |  |
| <sup>2</sup> Pearson's Chi-squared test; Wilcoxon rank sum test |  |  |  |  |

A total of 191 individuals had available data on plasma p-tau measurements and Aβ-PET while a subset of 142 had measurements from the Neurology 4-Plex E assay (NfL, GFAP, Aβ42, Aβ40) and for Aβ-PET. Of these, 96 individuals were used for training and 95 for testing for the p-tau assays. For the 4-plex assays, 72 individuals were used for training and 70 for testing. Likewise, 193 individuals had available data on plasma p-tau measurements and CT-based MRI, 97 of which were used for training and 96 for testing. A subset of 144 participants had information for the 4-plex assays and CT-based MRI data, 72 of which were used for training and 72 for testing.

### 3.2 Association of plasma biomarkers with amyloid pathology (A status)

Several plasma biomarkers showed statistically significant differences between A+ and A- groups (Table 1 and Figure 1). The ALZpath p-tau217 assay had higher median levels (p<0.001) in the A+ (0.51 pg/mL, IQR: [0.40, 0.80] pg/mL) versus the A- group (0.24 pg/mL, IQR: [0.17, 0.31] pg/mL). The same trend was seen for the Janssen p-tau217+ levels (A+: 0.070 IQR: [0.049, 0.105] pg/mL vs. A-: 0.033, IQR: [0.025, 0.044] pg/mL, p<0.001). Significant differences between the A+ and A- groups were also observed for p-tau181 (p=0.014), with median levels at 16 (IQR: [12, 23]) pg/mL and 12 (IQR: [9, 18]) pg/mL, respectively. Similarly, plasma GFAP levels were higher in the A+ (92, IQR: [79, 155] pg/mL) versus the A- (63, IQR: [46, 95] pg/mL) groups (p<0.001). The NfL assay had higher median levels in the A+ group (18; IQR: [11, 22] pg/mL) than the A- group (12, IQR: [8, 17] pg/mL) (p=0.003). The median plasma Aβ42/40 ratio was lower (p<0.001) in the A+ group (0.060, IQR: [0.056, 0.069]) compared with the A- group (0.074, IQR: [0.065, 0.083]), as expected. There was no significant median level difference for p-tau231 (p=0.472) between A+ (10, IQR: [6, 17] pg/mL) and A- (9, IQR: [6, 13] pg/mL) individuals.

**Figure 1.**
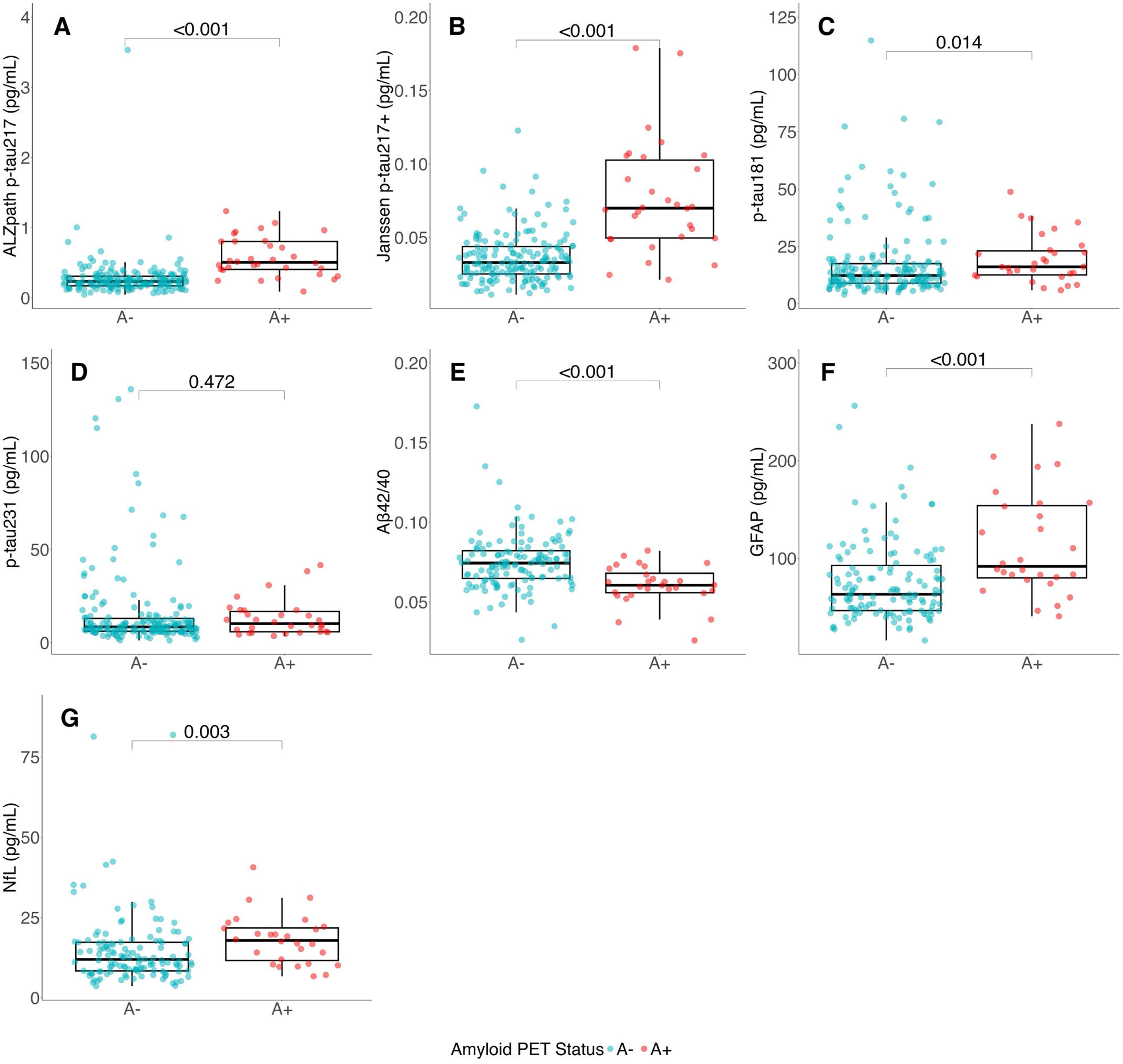
Plasma biomarker profiles according to dichotomized Aβ PET status. The boxplots with overlaid data points showed the distribution of plasma biomarkers in Aβ PET positive vs. negative groups: (A) ALZpath p-tau217, (B) Janssen p-tau217+, (C) p-tau181, (D) p- tau231, (E) Aβ42/40, (F) GFAP, and (G) NfL. The box represents the interquartile range, with the end points as the 25^th^ and 75^th^ percentiles, and the median line within the box. The whiskers are the most extreme non-outlier points, and any points beyond the whiskers are more than 1.5*IQR lower than Q1 or higher than Q3. P-values displayed are from the Wilcoxon rank sum test.

Among all plasma biomarkers, the p-tau217 assays had the largest fold changes when comparing A+ and A- groups, with Janssen having a fold change of 2.20 and ALZpath of 2.14. These were followed by NfL, GFAP, p-tau181, and p-tau231, with fold changes of 1.52, 1.50, 1.33, and 1.19, respectively. Aβ42/40 was lower in A+, at 0.81 times that of A-.

We next examined the strength of the correlation between plasma biomarkers and brain Aβ plaque burden, as assessed by PiB SUVR (Figure S1). All biomarkers showed significant association except for p-tau231 (p=0.125). Positive correlations were in the following decreasing order; Janssen p-tau217+ (R=0.38, p<0.001), GFAP (R=0.35, p<0.001), ALZpath p-tau217 (R=0.33, p<0.001), p-tau181 (R=0.16, p=0.024), and NfL (R=0.21, p=0.009). Aβ42/40 showed an inverse correlation, with R of -0.38, p<0.001.

### 3.3 Predictive accuracy of plasma biomarkers for Aβ PET status

For plasma biomarker-only models, we found that both p-tau217 assays showed a high predictive ability to distinguish the A+ from the A- participants (Figure 2A), with AUCs of 0.919 (95% CI = [0.859, 0.981]) for ALZpath and 0.914 (95% CI = [0.837, 0.992]) for Janssen assay. P-tau181 and p-tau231 were inferior to p-tau217, with AUCs of 0.676 (95% CI = [0.543, 0.808]) and 0.544 (95% CI = [0.366, 0.722]), respectively (Table S1).

**Figure 2.**
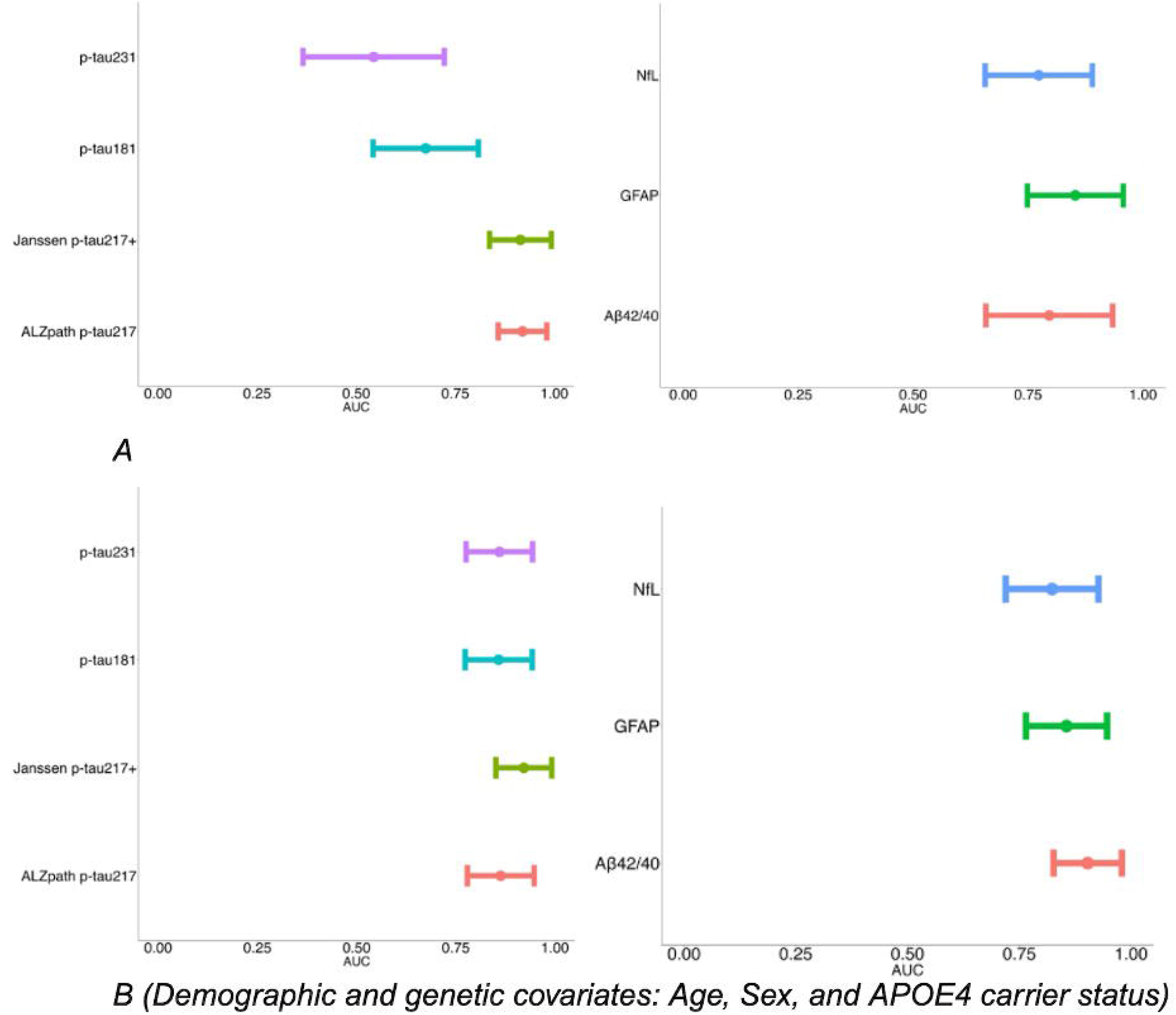
Forest plots indicating AUCs of plasma biomarkers for distinguishing Aβ PET status in participants. (A) AUCs based on biomarker-alone models, and (B) AUCs after adjusting for demographic and genetic variables (age, sex, and *APOE* ε4 carrier status). The center dots represent the point estimate of AUC values, and the lines represent the 95% CI.

We then utilized multivariable models to adjust demographic and genetic covariates (age, sex, and *APOE* ε4 carrier status) and compared the multivariable models to the corresponding biomarker-only models (Figure 2B). The inclusion of the covariates significantly increased the AUCs for p-tau181 to 0.858 (95% CI = [0.774, 0.942]) and the p-tau231 to 0.860 (95% CI = [0.776, 0.944]), with p values of 0.023 and 0.003, respectively (Figure S2). The AUC for Janssen p-tau217+ showed a slight but non-significant increase to 0.922 (95% CI = [0.851, 0.992]) (p = 0.813). ALZpath, on the other hand, showed a slight decrease of AUC to 0.863 (95% CI = 0.779, 0.948]). However, the difference was also not significant (p=0.275).

Within the 4-plex assays (Figure 2A), GFAP (AUC = 0.853, 95% CI = [0.749, 0.957]) had the highest AUC for differentiating A+ and A- participants, followed by Aβ42/40 (AUC = 0.796, 95% CI = [0.658, 0.934]) and NfL (AUC = 0.773, 95% CI = [0.657, 0.890]) (Table S1). Adding demographic covariates resulted in slightly higher AUCs for GFAP (AUC = 0.856, 95% CI = [0.765, 0.946]), NfL (AUC = 0.823, 95% CI = [0.719, 0.927]), and Aβ42/40 (AUC = 0.903, 95% CI = [0.826, 0.979]) (Figure 2B & Table S2). However, none of the increases were statistically significant (p>0.05).

### 3.4 Plasma biomarker association with neurodegeneration (N status)

When dichotomizing participants based on N status, only p-tau181 and p-tau231 showed significant differences, with p values of 0.011 and 0.029, respectively (Table 2, Figure 3). P- tau181 displayed the highest fold change increase, with N+ being 1.18 times that of N- individuals. The level of p-tau231 was 1.11 times higher in N+ compared to N-. P-tau217 from both ALZpath and Janssen, GFAP, and NfL levels also showed a slight increase in N+ compared to N-. However, none of these increases were statistically significant (Figure 3). The ALZpath (p=0.109) and Janssen (p=0.161) p-tau217 levels were 1.11 times higher in N+ compared to N-. The level of GFAP was 1.16 times higher in N+ compared to N- (p=0.517), and the level of NfL was 1.15 times higher in N+ compared to N- (p=0.281). Aβ42/40 ratio was 0.99 times lower in N+ compared to N-, but this was also statistically insignificant (p=0.591).

**Figure 3.**
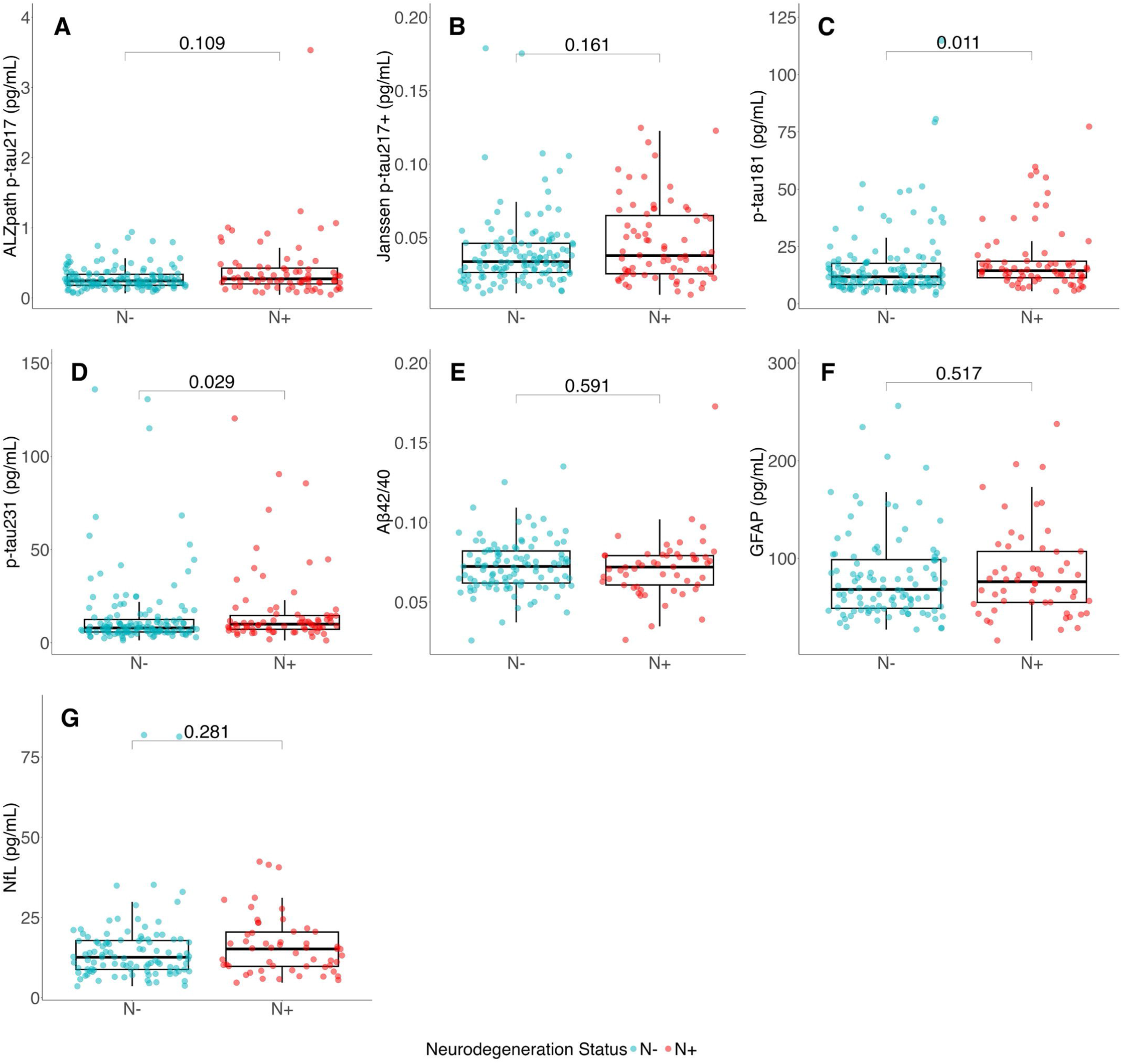
Plasma biomarker profiles according to neurodegeneration status. Shown are the boxplots with overlaid data points for the distribution of plasma biomarker levels in N positive and negative groups: (A) ALZpath p-tau217, (B) Janssen p-tau217+, (C) p-tau181, (D) p- tau231, (E) Aβ42/40, (F) GFAP, and (G) NfL. The box represents the interquartile range, with the end points as the 25^th^ and 75^th^ percentiles, and the median line within the box. The whiskers are the most extreme non-outlier points, and any points beyond the whiskers are more than 1.5*IQR lower than Q1 or higher than Q3. P-values displayed are from the Wilcoxon rank sum test.

We also evaluated the association of plasma biomarkers with the continuous measure of cortical thickness. Both p-tau181 and p-tau231 showed a strong inverse relationship with cortical thickness, with Spearman correlation coefficient of -0.24 (p<0.001) and -0.19 (p=0.005), respectively. Janssen p-tau217+ also showed a marginally significant inverse relationship, with R of -0.14 (p=0.047) (Figure S2). The remaining biomarkers, including ALZpath p-tau217, GFAP, NfL, and Aβ42/40, showed no significant association with cortical thickness (all p>0.05).

### 3.5 Predictive accuracy of plasma biomarkers for N status

All p-tau assays displayed low predictive capabilities in differentiating N+ from N- participants (Figure 4A and Table S3). Specifically, p-tau181 (AUC = 0.615, 95% CI = [0.494, 0.737]) had the highest AUC followed by ALZpath p-tau217 (AUC = 0.601, 95% CI = [0.478, 0.724]), p-tau231 (AUC = 0.585, 95% CI = [0.460, 0.710]), and Janssen p-tau217+ (AUC = 0.558, 95% CI = [0.422, 0.693]). Including demographic and genetic covariates (age, sex, race, and *APOE* ε4 status) increased the AUCs of p-tau. However, none of the increases were statistically significant (all p>0.05) (Figure 4B and S4).

**Figure 4.**
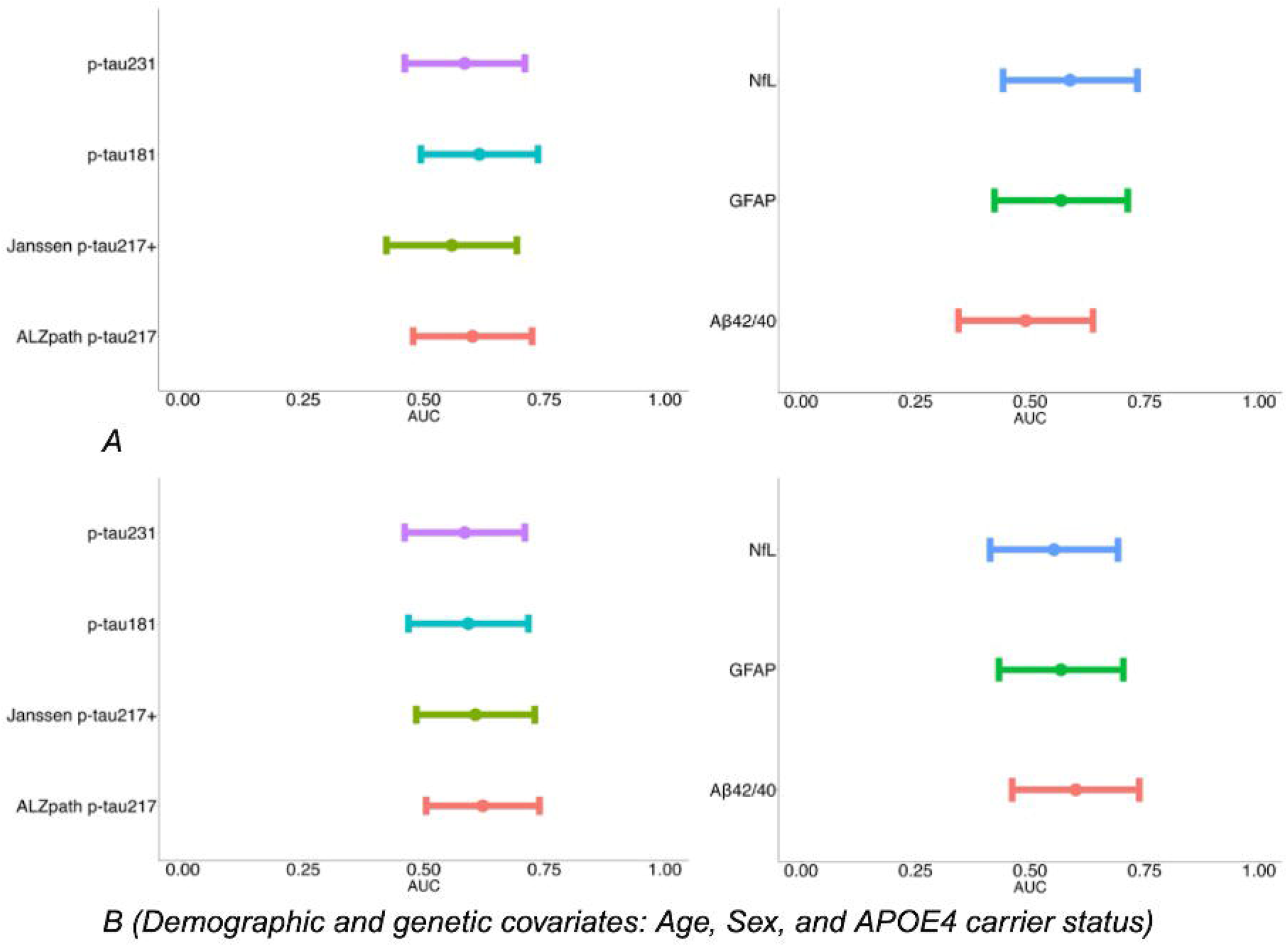
Forest plots illustrating the predictive performance of plasma biomarkers on neurodegeneration status. The center dots represent the point estimate of AUC values, and the lines represent the 95% CI. (A) AUCs based on biomarker-alone models, and (B) AUCs after adjusting for demographic and genetic variables (age, sex, and *APOE* ε4 carrier status).

In terms of the 4-plex assays, NfL (AUC = 0.587, 95% CI = [0.441, 0.734]) had a slightly higher AUC compared to GFAP (AUC = 0.568, 95% CI = [0.422, 0.713]) and Aβ42/40 (AUC = 0.490, 95% CI = [0.344, 0.637]) (Figure 4B and Table S3). Adding the same covariates as above to the models resulted in little to no change in their predictive performance for N status, with AUCs of 0.552 for NfL (95% CI = [0.413, 0.692]), 0.568 for GFAP (95% CI = [0.432, 0.703]), and 0.600 for Aβ42/40 (95% CI = [0.461, 0.739]) (Figure 4B and Table S4).

### 3.6 Plasma biomarker profiles in combined A and N status groups

We next evaluated whether the observed significance between dichotomized A status depends on N status, and vice versa, by categorizing participants into four groups according to the combined A and N statuses. Figure 5 shows the results of the statistical comparisons among groups, and Table S5 shows the demographic distribution. All except p-tau231 showed overall significance among the four groups, with Kruskal-Wallis p values <0.001 for ALZpath p- tau217, Janssen p-tau217+, GFAP, and Aβ42/40, and p values of 0.012 for p-tau181 and 0.025 for NfL. In addition, p-tau231 and NfL saw no differences across any pairwise comparisons among the four groups after the Bonferroni correction. Also, none of the biomarkers had statistically significant differences between the N- and N+ groups after controlling for the A status (A-N- vs. A-N+, and A+N- vs. A+N+).

**Figure 5.**
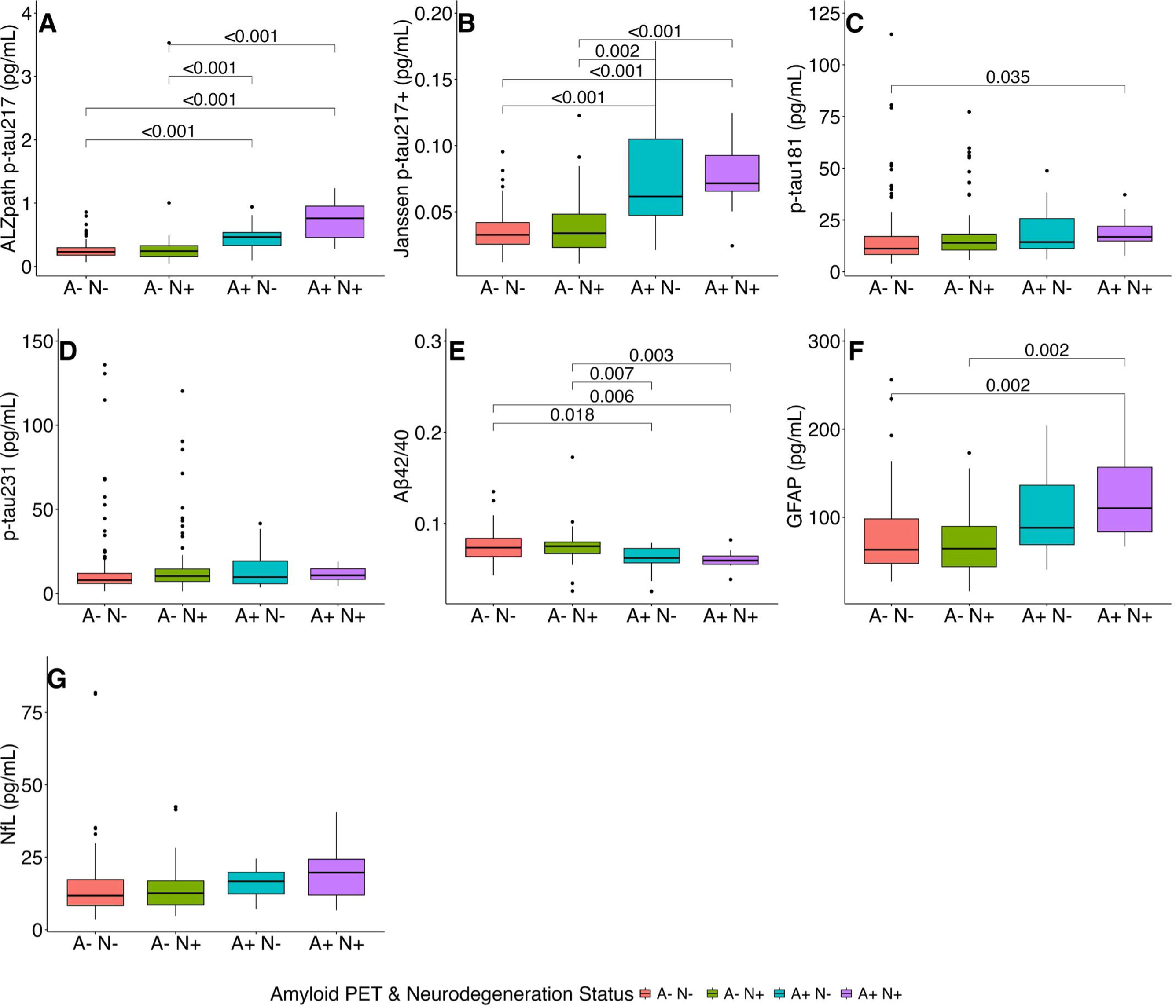
Plasma biomarker profiles according to combined A and N statuses. Shown are boxplot distributions of plasma biomarker levels among the four groups categorized by A and N status. The box represents the interquartile range, with the end points as the 25^th^ and 75^th^ percentiles, and the median line within the box. The whiskers are the most extreme non-outlier points, and any points beyond the whiskers are more than 1.5*IQR lower than Q1 or higher than Q3. Significant p-values are shown from Wilcoxon rank sum tests with a Bonferroni correction. (A) ALZpath p-tau217, (B) Janssen p-tau217+, (C) p-tau181, (D) p-tau231, (E) Aβ42/40, (F) GFAP, and (G) NfL.

ALZpath p-tau217, Janssen p-tau217+, and Aβ42/40 showed significant differences between A- and A+, regardless of N status. However, all three biomarkers showed larger A+/A- fold change among N+ participants, with fold changes of 3.21 (p<0.001) for ALZpath p-tau217, 2.44 (p<0.001) for Janssen p-tau217+, and 0.79 (p=0.002) for Aβ42/40 among N+ participants, compared to 1.92 (p<0.001), 2.05 (p<0.001), and 0.845 (p=0.018) in N- participants. In contrast, GFAP only showed significant differences among N+ groups (A-N+ vs. A+N+). Its level was 1.76 times higher in A+ compared to A- among N+ participants (p=0.002), compared to a fold change of 1.41 in N- participants (p=0.28).

### 3.7 Impact of self-identified race on the link between plasma biomarkers and Aβ PET status

When grouped by self-identified race, there were significant differences in several plasma biomarkers. The level of p-tau231 was 1.33 times higher in those who identified as B/AA compared to NHW (p<0.001), followed by p-tau181 with levels 1.15 times higher in B/AA compared to NHW (p=0.018). The median fold change comparing B/AA to NHW for Janssen p-tau217+ was 0.87 (p=0.002), and the ALZpath p-tau217 levels were 0.81 times lower in those who identified as B/AA compared to NHW (p<0.001). The median level for GFAP was 0.78 times lower for B/AA compared with NHW (p=0.039). Aβ42/40 levels were 1.03 times higher in B/AA vs. NHW, but this was not significantly different (p=0.354). NfL median levels were 0.913 times lower for those who identified as B/AA compared to NHW (p=0.066).

Cohort characteristics, separated by self-identified race and Aβ-PET status, are shown in Supplementary Table S6. As shown in the table, several plasma biomarkers exhibited racial-dependent associations with Aβ-PET status. For example, while plasma p-tau217 (measured by both Janssen and ALZpath) and GFAP showed a statistically significant difference between A+ and A- regardless of self-identified race, the fold change increases were larger in B/AA participants vs. NHW participants. Janssen p-tau217+ had a fold change of 2.29 versus 1.91 (A+/A-) for B/AA and NHW, respectively, but the interaction between race and Janssen p- tau217+ in the logistic regression model was not significant (p=0.135). ALZpath p-tau217 showed a fold change of 2.57 for B/AA compared to 1.97 for NHW, and the interaction with race was significant (p=0.024). Similarly, GFAP showed a fold change of 1.89 in B/AA compared to 1.36 in NHW, albeit there was no significant interaction (p=0.330). Aβ42/40 showed significant differences between A+ and A- regardless of self-identified race, but the fold change of 0.80 was greater for those who identified as white compared to the fold change of 0.82 within the B/AA group. The logistic regression interaction term between race and Aβ42/40 was insignificant (p=0.784).

For p-tau181 and p-tau231, significant differences between the A+ and A- groups were recorded only in the NHW group, not in the B/AA group (Figure 6). Specifically, p-tau181 had a fold change of 1.65 in B/AA versus 1.51 in NHW. For p-tau231, the fold change was 1.47 for B/AA versus 1.35 for NHW. In contrast, NfL exhibited a significant difference between A+ and A- only in B/AA participants, with a fold change of 2.15 in B/AA and 1.39 in NHW. None of the logistic regression interaction terms between race and p-tau181, p-tau231, or NfL were significant when predicting A status, with p-values of 0.171, 0.269, and 0.322, respectively.

**Figure 6.**
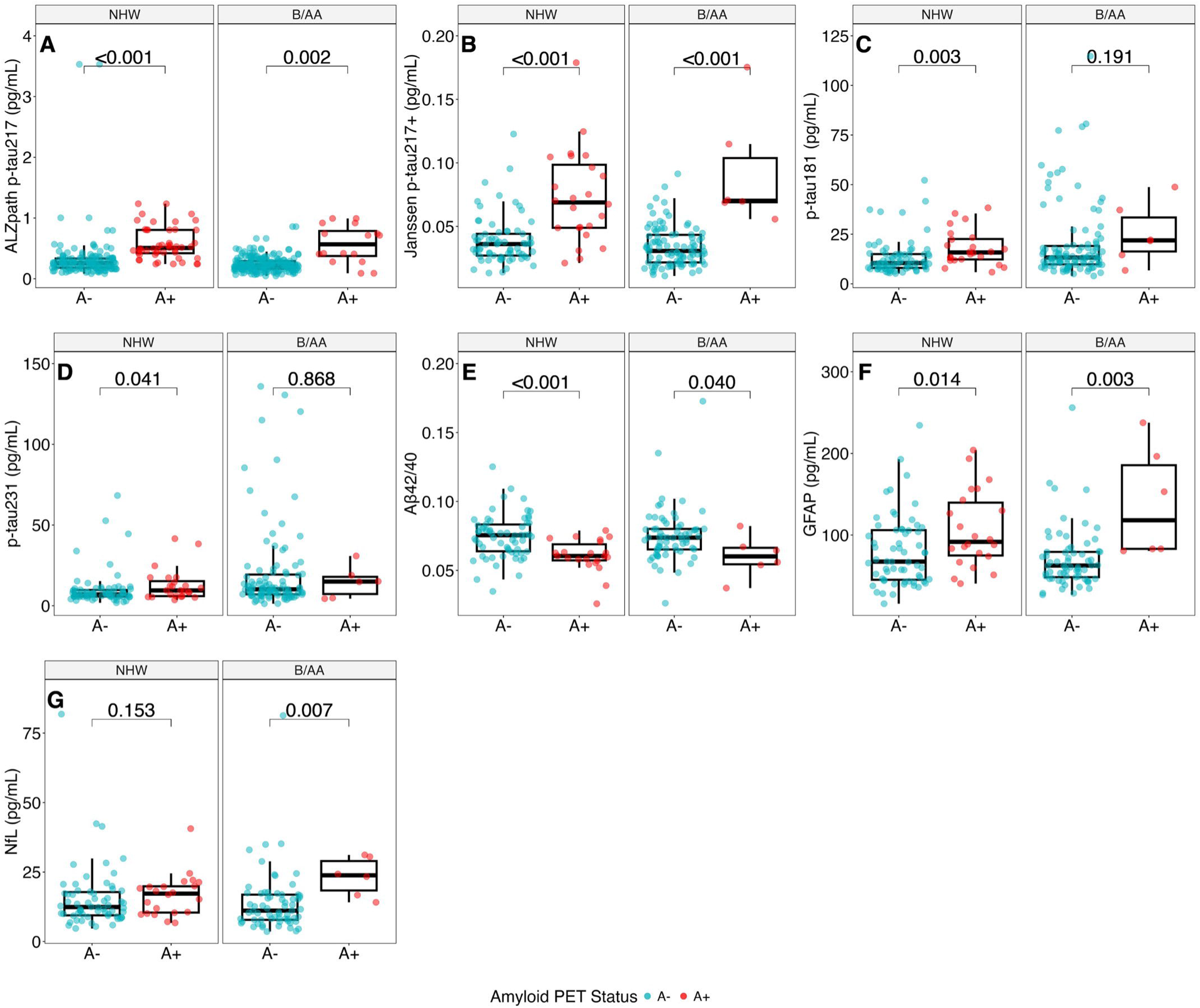
Plasma biomarker levels according to Aβ PET status classified based on self-identified Black/African American (B/AA) vs. non-Hispanic White (NHW) participants. (A) ALZpath p-tau217, (B) Janssen p-tau217+, (C) p-tau181, (D) p-tau231, (E) Aβ42/40, (F) GFAP, and (G) NfL. Statistical comparisons were made using the Wilcoxon rank sum test for self-identified racial groups. P-values are shown at the top of each plot, underneath the NHW or B/AA title. The box represents the interquartile range, with the end points as the 25^th^ and 75^th^ percentiles, and the median line within the box. The whiskers are the most extreme non-outlier points, and any points beyond the whiskers are more than 1.5*IQR lower than Q1 or higher than Q3. See Supplementary Figure S3 and Table S6 for further analysis of these results.

We evaluated whether there were any racial differences in the association of plasma biomarkers with amyloid burden assessed by global PiB SUVR (Figure S3). Significant racial differences, as determined by the significance of the interaction term between race and biomarkers in the linear regression models, were observed for p-tau217 measured with Janssen assay, p-tau181, and Aβ42/40, with p values of <0.001, 0.005, and 0.004, respectively. NHW was associated with a steeper incline in global PiB SUVR with increased Janssen p-tau217+ (0.086 for those who self-identified as B/AA vs. 0.208 increase per standard deviation in NHW). A similar steeper incline was observed for p-tau181, with an increase from 0.003 for B/AA to 0.134 for NHW in global Aβ PiB SUVR per standard deviation increase in p-tau181. Aβ42/40, on the other hand, showed a much steeper decrease in NHW, with a 0.027 decrease in PiB SUVR per standard deviation increase in Aβ42/40 in B/AA, compared to a 0.150 decrease for those who identified as NHW. The ALZpath p-tau217 (p=0.435), p-tau231 (p=0.109), GFAP (p=0.247), and NfL (p=0.979) biomarker assays did not show a significant racial impact on their association with global PiB SUVR and the assays.

## 4. DISCUSSION

The use of plasma biomarkers in blood tests for Aβ pathology is critical in the effort to independently detect AD preclinically and symptomatically in an effective, efficient, and less invasive manner than using CSF and neuroimaging methods. While many plasma biomarkers have been shown to be related to Aβ pathology, it is still unclear which amalgamations are superior in predicting A status and N status in a heterogeneous population.

We evaluated the classification accuracies of several plasma biomarkers for brain Aβ pathology and neurodegeneration in a racially diverse community cohort. While there have been a number of studies demonstrating high performances of plasma biomarkers, especially p-tau217, to identify abnormal brain Aβ PET, the vast majority of those studies lack diverse representation and are heavily comprised of self-identified NHW participants^39^. Hence, there is a need for studies focused on widening the participation of diverse populations in biomarker studies.

Plasma p-tau217 assays from Janssen and ALZpath demonstrated superior diagnostic accuracy in identifying Aβ pathology status, followed by GFAP, Aβ42/40, and p-tau181. These findings agree with those from several other cohorts^40^ including community/population-based ones^6,40–42^, establishing that the value of these biomarkers might be applicable to wider populations. Furthermore, adjusting the analysis for demographic covariates did not provide much improvement to the AUCs, indicating that the biomarkers themselves can perform well unaided. In a cohort of only 15% A+ individuals and substantially cognitively normal participants, the plasma p-tau217 assays had the best performance as triaging tests.

For plasma biomarker association with cortical thickness, the lack of significant AUC values with N status suggests that these biomarkers individually may not be adequate predictors of N status, thus calling for novel AD-type N status markers anticipated in future analysis. A potentially useful marker might be brain-derived tau, which has been shown to be a plasma biomarker that associates with AD-specific neurodegenerative features^43,44^.

Results based on comparing plasma biomarker profiles among groups stratified by A and N statuses indicated that, compared to the A-N- group, the A+N+ group showed the largest differences in p-tau181, p-tau217, GFAP, and Aβ42/40 levels, followed closely by the A+N- group. These findings suggest that Aβ pathology is the primary driver of increased plasma biomarker levels, with neurodegeneration contributing further but with more subtle abnormalities, particularly in the presence of brain Aβ pathology. These results align with published findings that core AD biomarkers, particularly p-tau181 and p-tau217, are associated with the earlier phases of AD pathophysiology^40,45–48^.

Racial differences in plasma biomarkers for AD have been increasingly recognized, though the underlying causes remain complex and multifactorial. Studies have reported that B/AA individuals tend to show lower levels of p-tau biomarkers (such as p-tau181 and p-tau217) and reduced tau PET binding compared to NHW individuals, even when matched for disease stage^49^. Findings regarding Aβ42/40 ratios are more variable, with some studies indicating higher or similar ratios in B/AA individuals, while others, including ours, observe the opposite^50^. NfL and GFAP levels may also differ by race, but evidence remains limited ^51^. These differences may not reflect innate biological variation, but rather be influenced by social determinants of health, including access to healthcare, comorbid conditions (e.g., hypertension, diabetes), chronic stress, and structural racism. Furthermore, we are not certain what exactly is causing the observed differences between p-tau217 assays Janssen and ALZpath in predicting the probability of amyloid burden based on biomarker and global PiB SUVR in the linear regression nor what is accounting for the difference between the p-tau217 assays in the logistic regression model. Perhaps the differences can be accounted for by the varying affinities to different tau isoforms; Janssen p-tau217+ has been shown to have high affinity when there is additional phosphorylation at 212, but further research is needed to explore these varying effects^52^. Consequently, biomarker thresholds and interpretations need to be contextualized within diverse populations to ensure accurate and equitable diagnosis and treatment.

We observed lower levels of plasma p-tau biomarkers in B/AA individuals, in agreement with some previous literature (Figure 6)^41^. The fold changes were higher in B/AA for all biomarkers. However, we observed an unexpected lower Aβ42/40 ratio in B/AA compared to NHW individuals, opposing several prior publications that found higher or similar Aβ42/40 ratios in B/AA and NHW individuals^41,53^. While the reasons accounting for our findings cannot be ascertained currently, they could perhaps be a factor of the social determinants of health, specifically as it relates to comorbidities and health conditions. The discrepancies between our findings and existing literature may reflect underlying social determinants of health such as differential access to care, comorbidity profiles (e.g., vascular risk), educational opportunities, and chronic stress exposure, rather than innate racial differences^54^.

A significant strength of this study is the diversity of the cohort. Most studies examining the association between plasma biomarkers and AD pathology have predominantly included non-Hispanic White participants in their cohorts. In contrast, our study included almost equal representation of Black/African American and non-Hispanic White volunteers. This allowed us to examine the racial impact on the AT(N) or AT/N/I/V/S frameworks for their real-world application. Additional advantages of this study include correlating biomarkers to both PET-PiB imaging and MRI-based cortical thickness, and the evaluation of multiple biomarkers that reflect different pathophysiological processes in AD. Our study is limited by the lack of imaging information for tau status, and also, only two self-identified racial groups were included – Black/African American and non-Hispanic White.

Together, this study evaluated the clinical performance of the plasma biomarkers in identifying neuroimaging-based A and N statuses. Our study indicated that p-tau217, measured by both the Janssen and ALZpath assays, was the best predictor of Aβ-PET pathology for triaging purposes. No substantial improvement was observed with the inclusion of covariates (age, sex, and *APOE* ε4 carrier status). P-tau181 and p-tau231, but not p-tau217, showed significant correlation with neurodegeneration status (based on cortical thickness). Self-identified racial identity significantly influenced the association between p-tau217 (Janssen), p-tau181, Aβ42/40, and Aβ plaque burden measured by PiB SUVR. This deserves further attention to ascertain the widespread nature of these findings and their root causes.

## Data Availability

All data produced in the present work are contained in the manuscript.

## Acknowledgements

We thank the HCP study participants and their families and caregivers.

## ACKNOWLEDGEMENTS/CONFLICTS/FUNDING SOURCES/CONSENT STATEMENT

### Consent statement

All participants provided written consent, and the University of Pittsburgh Institutional Review Board approved the study.

