## Supplemental for "Plasma biomarkers, brain amyloid pathology, and cortical thickness in a diverse middle-aged community cohort: the HCP-CoBRA study"

**Supplementary file**

**Figure S1. Scatterplot distribution of plasma biomarkers (y-axis) and global PiB SUVR (X-axis).**

**Table S1. Predictive metrics and associated AUCs for predicting Aβ PET status using the biomarker only models.**

**Table S2. Predictive metrics and associated AUCs for predicting Aβ PET status using the biomarker and demographic adjusted models.**

**Figure S2. Correlation of plasma biomarker concentrations with cortical thickness.**

**Table S3. Predictive metrics and associated AUCs for predicting Neurodegeneration status using the biomarker only models.**

**Table S4. ROC curves and associated AUCs for predicting Neurodegeneration status using the biomarker and demographic adjusted models.**

**Figure S3. Correlation of standardized plasma biomarkers with Aβ PET uptake according to self-identified Black/African American (B/AA) vs. non-Hispanic White (NHW) racial groups.**

**Table S5. Participant characteristics of the HCP cohort according to A and N combined statuses**

**Table S6. Cohort charactersitics according to self-identified race on plasma biomarker association with Aβ PET uptake.**

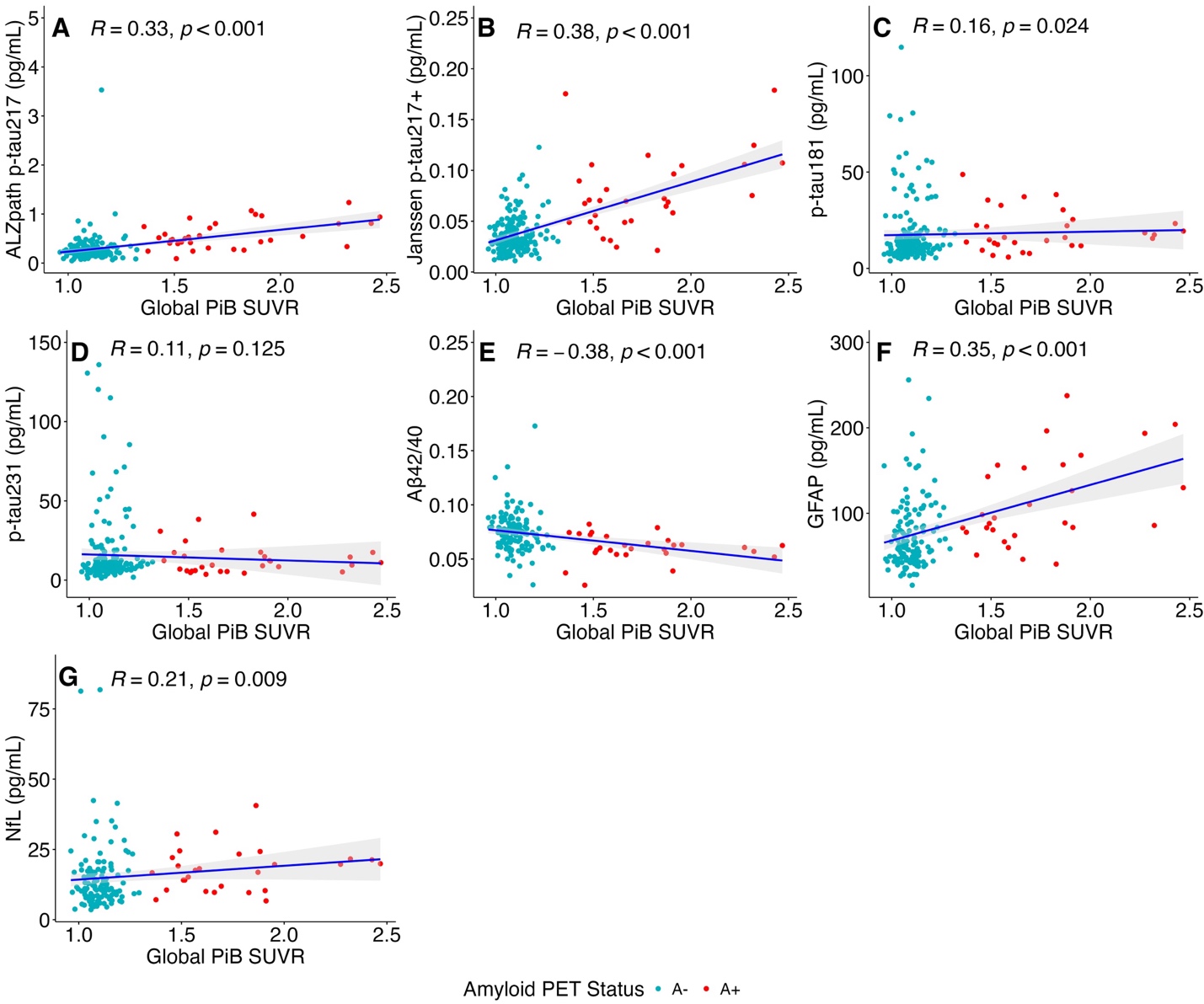

**Figure S1. Scatterplot distribution of plasma biomarkers (y-axis) and global PiB SUVR (X-axis).** The data points were color-coded according to the Aβ PET status, with blue dots for Aβ PET-negative and red dots for Aβ PET-positive. Correlation coefficients and corresponding p values were determined using Spearman correlation. The blue line indicates the best-fit linear regression line, and the grey zones represent the 95% confidence interval of the regression lines. A) ALZpath p-tau217, (B) Janssen p-tau217+, (C) p-tau181, (D) p-tau231, (E) Aβ42/40, (F) GFAP, and (G) NfL.  **Table S1. Performance characteristics for predicting Aβ PET status using the biomarker-only models.**

|  | **Sensitivity** | **Specificity** | **PPV** | **NPV** | **AUC** |
| --- | --- | --- | --- | --- | --- |
| **ALZpath p-tau217** | 0.857 | 0.889 | 0.571 | 0.973 | 0.919 |
| **Janssen p-tau217+** | 0.857 | 0.852 | 0.500 | 0.972 | 0.915 |
| **p-tau181** | 0.929 | 0.469 | 0.232 | 0.974 | 0.676 |
| **p-tau231** | 0.500 | 0.284 | 0.108 | 0.767 | 0.544 |
| **Aß42/40** | 0.769 | 0.790 | 0.455 | 0.938 | 0.796 |
| **GFAP** | 0.923 | 0.702 | 0.414 | 0.976 | 0.853 |
| **NfL** | 0.846 | 0.684 | 0.379 | 0.951 | 0.773 |

Sensitivity, specificity, positive predictive value (PPV), and negative predictive value (NPV) were determined with optimal Youden’s index.

**Table S2. Performance metrics for predicting Aβ PET status using covariate-adjusted models.**

|  | **Sensitivity** | **Specificity** | **PPV** | **NPV** | **AUC** |
| --- | --- | --- | --- | --- | --- |
| **ALZpath p-tau217** | 0.857 | 0.765 | 0.387 | 0.969 | 0.863 |
| **Janssen p-tau217+** | 0.714 | 0.988 | 0.909 | 0.952 | 0.922 |
| **P-tau181** | 1.000 | 0.617 | 0.311 | 1.000 | 0.858 |
| **P-tau231** | 0.857 | 0.765 | 0.387 | 0.969 | 0.860 |
| **Aß42/40** | 0.846 | 0.842 | 0.550 | 0.960 | 0.901 |
| **GFAP** | 1.000 | 0.649 | 0.394 | 1.000 | 0.856 |
| **NfL** | 0.923 | 0.684 | 0.400 | 0.975 | 0.823 |

Demographic and genetic covariates include age, sex, and *APOE* ε4 carrier status*.* Sensitivity, specificity, positive predictive value (PPV), and negative predictive value (NPV) were determined with optimal Youden’s index.

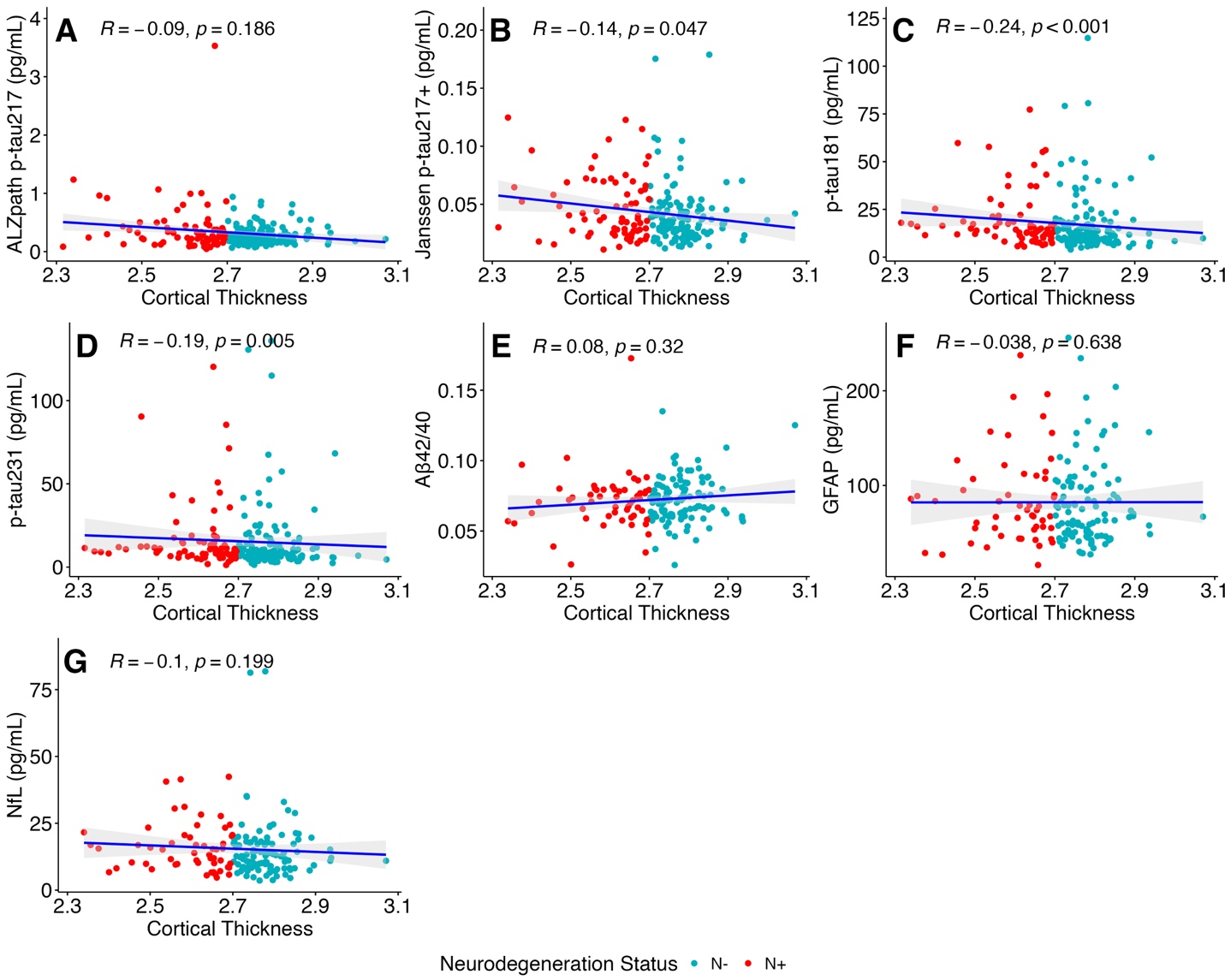

**Figure S2. Correlation of plasma biomarker levels with cortical thickness.** Correlation coefficients and their corresponding p values were determined using Spearman correlation. Blue lines and grey zones indicate the best-fit linear regression line and 95% CI, respectively. Data points were color-coded according to neurodegeneration status, with blue for N-, and red for N+. (A) ALZpath p-tau217, (B) Janssen p-tau217+, (C) p-tau181, (D) p-tau231, (E) Aβ42/40, (F) GFAP, and (G) NfL.

**Table S3. Performance metrics for predicting neurodegeneration status using the biomarker-only models.**

|  | **Sensitivity** | **Specificity** | **PPV** | **NPV** | **AUC** |
| --- | --- | --- | --- | --- | --- |
| **ALZpath p-tau217** | 0.471 | 0.758 | 0.516 | 0.723 | 0.601 |
| **Janssen p-tau217+** | 0.412 | 0.887 | 0.667 | 0.733 | 0.558 |
| **p-tau181** | 0.471 | 0.807 | 0.571 | 0.735 | 0.615 |
| **p-tau231** | 0.500 | 0.242 | 0.266 | 0.469 | 0.585 |
| **Aß42/40** | 0.440 | 0.447 | 0.297 | 0.600 | 0.490 |
| **GFAP** | 0.600 | 0.596 | 0.441 | 0.737 | 0.568 |
| **NfL** | 0.560 | 0.660 | 0.467 | 0.738 | 0.587 |

Sensitivity, specificity, positive predictive value (PPV), and negative predictive value (NPV) were determined with optimal Youden’s index.

**Table S4. Performance metrics for predicting neurodegeneration status using covariate-adjusted models.**

|  | **Sensitivity** | **Specificity** | **PPV** | **NPV** | **AUC** |
| --- | --- | --- | --- | --- | --- |
| **ALZpath p-tau217** | 0.941 | 0.323 | 0.432 | 0.909 | 0.622 |
| **Janssen p-tau217+** | 0.559 | 0.677 | 0.487 | 0.737 | 0.607 |
| **p-tau181** | 0.441 | 0.823 | 0.577 | 0.729 | 0.593 |
| **p-tau231** | 0.500 | 0.242 | 0.266 | 0.469 | 0.585 |
| **Aß42/40** | 0.160 | 0.638 | 0.190 | 0.588 | 0.600 |
| **GFAP** | 0.320 | 0.468 | 0.242 | 0.564 | 0.568 |
| **NfL** | 0.520 | 0.298 | 0.283 | 0.539 | 0.552 |

Demographic and genetic covariates include age, sex, and *APOE* ε4 carrier status. Sensitivity, specificity, positive predictive value (PPV), and negative predictive value (NPV) were determined with optimal Youden’s index.

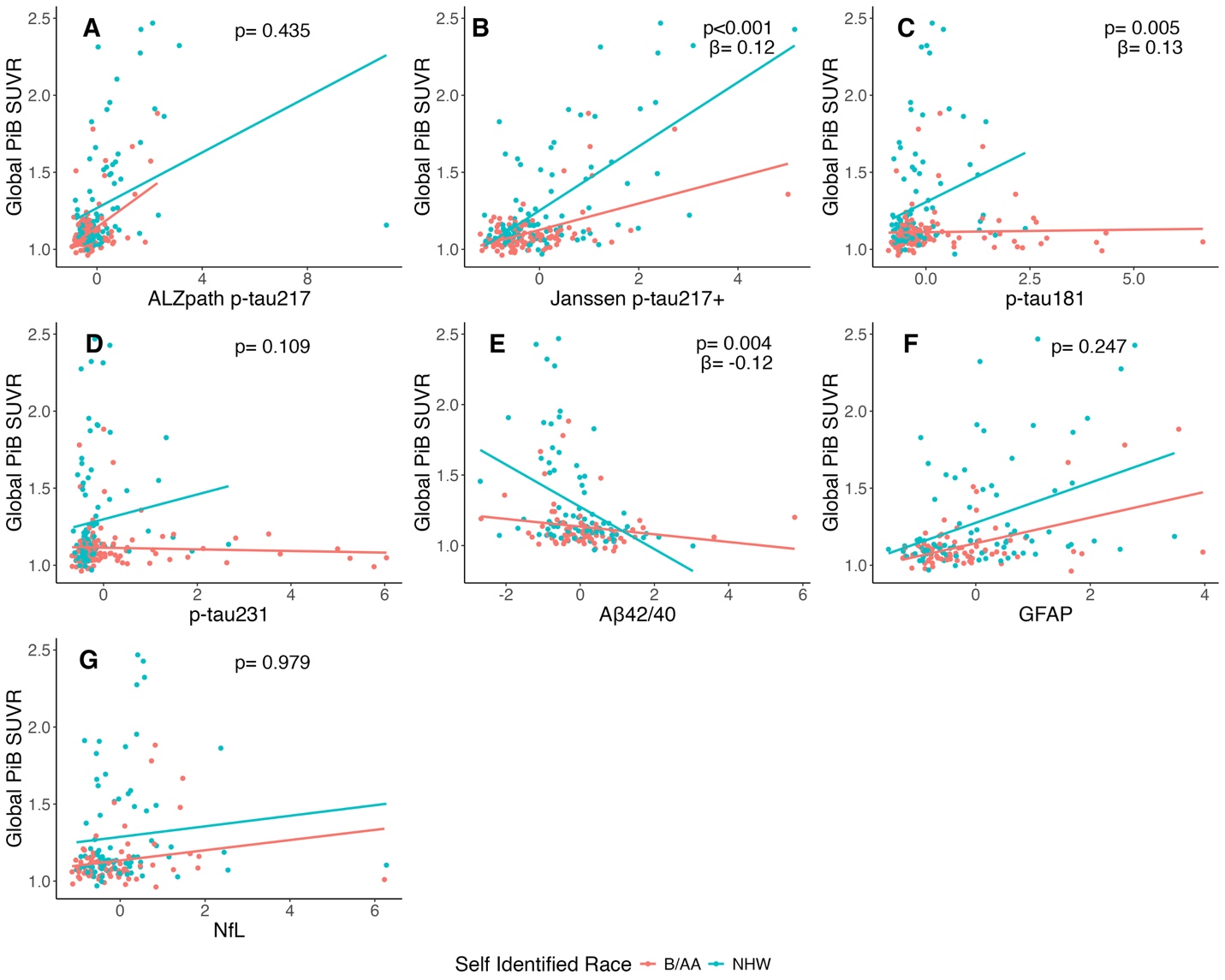

**Figure S3. Correlation of standardized plasma biomarkers with Aβ PET uptake according to self-identified Black/African American (B/AA) vs. non-Hispanic White (NHW) racial groups.** P-values and β estimates (B/AA as reference) are from the interaction term between self-identified race and biomarkers in a regression model to predict Global PiB SUVR. Blue lines and dots represent self-identified NHW participants. The red lines and dots represent self-identified B/AA participants. Only β estimates with a significant p-value are displayed. (A) ALZpath p-tau217, (B) Janssen p-tau217+, (C) p-tau181, (D) p-tau231, (E) Aβ42/40, (F) GFAP, and (G) NfL.

**Table S5. Participant characteristics according to the combined A and N statuses**

| **Table S5** | **A-N-**, N = 123^1^ | **A-N+**, N = 62^1^ | **A+N-**, N = 18^1^ | **A+N+**, N = 15^1^ | **p-value**^2^ |
| --- | --- | --- | --- | --- | --- |
| **Sex** |  |  |  |  | 0.006 |
| Female | 79 (64%) | 48 (77%) | 9 (50%) | 5 (33%) |  |
| Male | 44 (36%) | 14 (23%) | 9 (50%) | 10 (67%) |  |
| **Age(years)** | 60 (56, 70) | 60 (55, 65) | 70 (66, 72) | 73 (71, 81) | <0.001 |
| **Self-identified race** |  |  |  |  | <0.001 |
| Black/African American | 66 (54%) | 42 (68%) | 3 (17%) | 5 (33%) |  |
| Non-Hispanic White | 57 (46%) | 20 (32%) | 15 (83%) | 10 (67%) |  |
| ***APOE4*** |  |  |  |  | 0.060 |
| Non-carrier | 90 (73%) | 41 (66%) | 14 (78%) | 6 (40%) |  |
| Carrier | 33 (27%) | 21 (34%) | 4 (22%) | 9 (60%) |  |
| **ALZpath p-tau 217 (pg/mL)** | 0.23 (0.18, 0.29) | 0.24 (0.16, 0.33) | 0.46 (0.31, 0.54) | 0.76 (0.43, 0.96) | <0.001 |
| (Missing) | 2 | 1 | 0 | 1 |  |
| **Janssen p-tau 217+ (pg/mL)** | 0.033 (0.026, 0.042) | 0.034 (0.023, 0.048) | 0.062 (0.046, 0.105) | 0.072 (0.065, 0.096) | <0.001 |
| (Missing) | 14 | 5 | 2 | 1 |  |
| **p-tau 181 (pg/mL)** | 11 (8, 17) | 14 (10, 18) | 14 (11, 28) | 17 (15, 22) | 0.012 |
| (Missing) | 4 | 3 | 2 | 1 |  |
| **p-tau 231 (pg/mL)** | 8 (6, 12) | 10 (7, 15) | 10 (6, 21) | 11 (8, 15) | 0.168 |
| (Missing) | 5 | 2 | 2 | 1 |  |
| **Aβ 40 (pg/mL)** | 69 (53, 80) | 68 (53, 78) | 60 (53, 85) | 65 (56, 76) | >0.900 |
| (Missing) | 36 | 22 | 3 | 2 |  |
| **Aβ 42 (pg/mL)** | 4.88 (3.90, 5.79) | 4.93 (3.92, 5.44) | 3.74 (3.16, 5.19) | 3.75 (3.51, 4.91) | 0.10 |
| (Missing) | 36 | 22 | 3 | 2 |  |
| **Aβ 42/40 ratio** | 0.074 (0.064, 0.084) | 0.075 (0.067, 0.080) | 0.062 (0.056, 0.073) | 0.060 (0.055, 0.064) | <0.001 |
| (Missing) | 36 | 22 | 3 | 2 |  |
| **GFAP (pg/mL)** | 63 (47, 99) | 64 (44, 90) | 88 (60, 143) | 110 (83, 157) | <0.001 |
| (Missing) | 36 | 22 | 3 | 2 |  |
| **NfL (pg/mL)** | 12 (8, 17) | 13 (8, 17) | 17 (11, 20) | 20 (12, 24) | 0.025 |
| (Missing) | 36 | 22 | 3 | 2 |  |
| **CT Composite Region** | 2.78 (2.74, 2.82) | 2.64 (2.57, 2.67) | 2.77 (2.73, 2.80) | 2.56 (2.42, 2.64) | <0.001 |
| **Global PiB SUVR Freesurfer** | 1.09 (1.05, 1.14) | 1.08 (1.05, 1.15) | 1.54 (1.48, 1.83) | 1.86 (1.62, 1.91) | <0.001 |
| ^1^n (%); Median (IQR) | | | | | |
| ^2^Pearson's Chi-squared test; Kruskal-Wallis rank sum test; Fisher's exact test | | | | | |

**Table S6. Cohort characteristics according to self-identified race and Aβ-PET positivity.**

| **Table S6** | **Black/African American** | | | **Non-Hispanic White** | | |
| --- | --- | --- | --- | --- | --- | --- |
|  | **Negative**, N = 108^1^ | **Positive**, N = 8^1^ | **p-value**^2^ | **Negative**, N = 77^1^ | **Positive**, N = 25^1^ | **p-value**^3^ |
| **Sex** |  |  | 0.053 |  |  | 0.083 |
| Female | 78 (72%) | 3 (38%) |  | 49 (64%) | 11 (44%) |  |
| Male | 30 (28%) | 5 (63%) |  | 28 (36%) | 14 (56%) |  |
| **Age at Enrollment (years)** | 59 (55, 65) | 73 (72, 75) | <0.001 | 64 (57, 71) | 71 (68, 79) | <0.001 |
| ***APOE4*** |  |  | 0.709 |  |  | 0.009 |
| Not Carrier | 68 (63%) | 6 (75%) |  | 63 (82%) | 14 (56%) |  |
| Carrier | 40 (37%) | 2 (25%) |  | 14 (18%) | 11 (44%) |  |
| **ALZpath p-tau 217 (pg/mL)** | 0.22 (0.16, 0.29) | 0.57 (0.34, 0.83) | 0.002 | 0.26 (0.18, 0.33) | 0.51 (0.41, 0.80) | <0.001 |
| (Missing) | 1 | 0 |  | 2 | 1 |  |
| **Janssen p-tau217+ (pg/mL)** | 0.031 (0.022, 0.043) | 0.070 (0.069, 0.115) | <0.001 | 0.036 (0.027, 0.044) | 0.069 (0.049, 0.101) | <0.001 |
| (Missing) | 11 | 2 |  | 8 | 1 |  |
| **p-tau181 (pg/mL)** | 13 (10, 20) | 22 (15, 37) | 0.191 | 10 (8, 15) | 16 (12, 23) | 0.003 |
| (Missing) | 5 | 2 |  | 2 | 1 |  |
| **p-tau231 (pg/mL)** | 10 (7, 20) | 15 (5, 19) | 0.868 | 7 (6, 10) | 9 (6, 16) | 0.041 |
| (Missing) | 5 | 2 |  | 2 | 1 |  |
| **Aβ40 (pg/mL)** | 65 (50, 78) | 81 (69, 94) | 0.105 | 72 (59, 83) | 61 (53, 70) | 0.109 |
| (Missing) | 43 | 2 |  | 15 | 3 |  |
| **Aβ42 (pg/mL)** | 4.61 (3.71, 5.33) | 5.30 (3.16, 6.21) | 0.657 | 5.11 (4.44, 6.02) | 3.74 (3.43, 4.64) | <0.001 |
| (Missing) | 43 | 2 |  | 15 | 3 |  |
| **Aβ42/40 ratio** | 0.074 (0.065, 0.080) | 0.060 (0.054, 0.067) | 0.040 | 0.075 (0.064, 0.083) | 0.060 (0.057, 0.071) | <0.001 |
| (Missing) | 43 | 2 |  | 15 | 3 |  |
| **GFAP (pg/mL)** | 63 (48, 79) | 118 (83, 196) | 0.003 | 68 (45, 106) | 92 (74, 143) | 0.014 |
| (Missing) | 43 | 2 |  | 15 | 3 |  |
| **NfL (pg/mL)** | 11 (8, 17) | 24 (17, 31) | 0.007 | 12 (9, 18) | 17 (10, 20) | 0.153 |
| (Missing) | 43 | 2 |  | 15 | 3 |  |
| **Global PiB SUVR Freesurfer** | 1.07 (1.04, 1.12) | 1.57 (1.49, 1.72) | <0.001 | 1.11 (1.07, 1.16) | 1.69 (1.53, 1.95) | <0.001 |
| **^1^n (%); Median (IQR)** | | | | | | |
| **^2^Fisher's exact test; Wilcoxon rank sum test** | | | | | | |
| **^3^Pearson's Chi-squared test; Wilcoxon rank sum test** | | | | | | |
